# Efficacy of platelet-rich products in preventing cerebrospinal fluid leakage following neurosurgical procedures: A systematic review and meta-analysis

**DOI:** 10.1101/2025.05.30.25328644

**Authors:** Jina Behjati, Mohammadreza Ghasemi, Shaghayegh Karami, Pooya Eini, Arman Keymoradzadeh, Mohammad Ansari, Hassan Reza Mohammadi, Mohsen Koosha, Saeed Oraee-Yazdani

**Author notes:** Corresponding authors at: Functional Neurosurgery Research Center, Shohada Tajrish Comprehensive Neurosurgical Center of Excellence, Shahid Beheshti University of Medical Sciences, Tehran, Iran (S. Oraee-Yazdani); Functional Neurosurgery Research Center, Shohada Tajrish Comprehensive Neurosurgical Center of Excellence, Shahid Beheshti University of Medical Sciences, Tehran, Iran (J. Behjati). These authors share senior authorship (Hassan Reza Mohammadi, Mohsen Koosha, Saeed Oraee-Yazdani).

## Abstract

**Background and objective:** Despite advancements in dural closure techniques and materials, cerebrospinal fluid (CSF) leakage remains a major issue for neurosurgeons. This systematic review assesses the efficacy of platelet-rich products in reducing post-operative CSF leakage.

**Method:** PubMed, Scopus, Web of Science, EMBASE, Clinicaltrials.gov, and Google Scholar were searched using the following keywords: “platelet-rich products”, “cerebrospinal fluid leak”, “cranial surgery”, and “spinal surgery”. Original studies using platelet-rich products to reduce CSF leaks were included. Meta-analysis used a random-effects model to calculate risk ratios and risk differences. The GRADE tool was used to assess the certainty of evidence. Statistical analyses were performed in STATA 17 and R.

**Results:** Eleven studies were included in the qualitative synthesis and five in the quantitative meta-analysis. Leukocyte-rich platelet fibrin (L-PRF) was the most commonly used product (k = 9). Most studies involved endoscopic endonasal skull base surgeries (K = 9). Meta-analysis demonstrated a non-significant effect in CSF leak prevention by relative risk analysis (RR: 1.03, 95% CI: 1.00-1.06, P = 0.06; I² = 0.01%), and risk difference analysis (RD: 0.07, 95% CI: −0.03 to 0.17, P = 0.18; I² = 72.57%). Qualitative analysis showed lower rates of meningitis and improved wound healing in intervention groups, with no serious adverse events attributed to platelet-rich products.

**Conclusion:** While platelet-rich products showed promise in preventing CSF leaks and reducing postoperative complications, their effectiveness was not statistically significant. Well-designed randomized controlled trials with larger populations are needed to establish their role in neurosurgical procedures.

## Introduction

Cerebrospinal fluid (CSF) leakage represents a common complication following cranial and spinal surgeries. The incidence varies based on procedure type and lesion location. In cranial procedures, skull base surgeries have an overall rate of 6.2%, with slight variations based on approach: 5.9% for anterior skull base, 6.4% for middle fossa, and 5.2% for transpetrosal approaches^1^. Posterior fossa craniotomies, however, have shown rates up to 32%^2^. When comparing supratentorial versus infratentorial approaches, CSF leaks occur in 2.9% and 5.8% of cases, respectively^1^. In spinal procedures, the rates vary by context: planned intradural surgeries have an 18% incidence, while incidental durotomies during other spinal procedures show rates ranging from 1% to 17%^2^. CSF leaks increase risks of infection, meningitis, hydrocephalus, and CSF fistula formation, which in turn leads to prolonged hospital stays, greater healthcare costs, and increased mortality^1,3^.

The cornerstone to prevent postoperative CSF leak is watertight dural closure through meticulous surgical reconstruction. However, primary dural closure alone has a CSF leak rate exceeding 40%^4^, necessitating supplementary reinforcement. These include reconstruction with various grafts and flaps (autologous, allogeneic, natural, and synthetic), which can be further enhanced with materials such as polymer hydrogels, liquid fibrin glues, and dural sealant patches^5,6^.

Recently platelet-rich products have emerged as a promising adjunct to reduce postoperative CSF leaks. These autologous products are derived from patients’ own blood through simple centrifugation, making them cost-effective and eliminating risks of immune reactions^7^. The resulting product is enriched with growth factors and immune cell products such as platelet-derived growth factor (PDGF), vascular endothelial growth factor (VEGF), epidermal growth factor (EGF), platelet-derived factor 4 (PF-4), and transforming growth factor-beta (TGF-β) which have demonstrated accelerated healing properties in various surgical fields ^8–10^. Among these products, leukocyte-platelet-rich fibrin (L-PRF) is the most widely used in neurosurgery. In addition to its regenerative benefits, leukocyte and platelet components help reduce the risk of infection^11^.

Despite the growing clinical use, the efficacy of platelet-rich products in preventing CSF leakage following neurosurgical procedures remains controversial. While some studies suggest reducing CSF leak rates with platelet-rich product applications^8^, others have failed to demonstrate significant benefits over conventional methods^11,12^.

In this systematic review and meta-analysis, we aim to evaluate the efficacy of platelet-rich products in reducing cerebrospinal fluid (CSF) leakage following cranial and spinal surgeries. We also provide an overview of their current applications in neurosurgery to establish their present role in clinical practice.

## Methods

This systematic review and meta-analysis was conducted following the Preferred Reporting Items for Systematic Reviews and Meta-Analysis (PRISMA) guidelines^13^ and was prospectively registered in the International Prospective Register of Systematic Reviews (PROSPERO) under protocol CRD42024619809. Our primary objective was to assess the effectiveness of platelet-rich products in preventing CSF leakage following cranial and spinal procedures.

### Search strategy

A comprehensive search was conducted across multiple databases, including PubMed, Scopus, Web of Science (WOS), EMBASE, Clinicaltrials.gov, and Google Scholar. The search included terms related to platelet-rich products and their variants, combined with terms for cranial and spinal surgical procedures. The initial search was completed on the 4th of December, 2024. An updated search was performed before manuscript submission to ensure inclusion of the most recent literature. Additionally, the reference lists of included studies were manually screened to identify relevant articles. The complete search strategy is detailed in the Supplementary Table 1.

### Eligibility criteria

We included studies that met the following criteria:

1. Patients of all ages undergoing cranial or spinal surgeries where platelet-rich products were used to prevent post-operative CSF leakage
2. Original research articles including randomized controlled trials (RCT), cohort studies, case-control studies, and case series.
3. Studies reporting CSF leakage and associated complications as outcomes

Studies were excluded if:

1. The study was published in a language other than English
2. The publication type was a case report, review, book chapter, editorial, comment, or conference abstract
3. The study was conducted on animal subjects or was in vitro
4. The study did not report CSF leak as an outcome

### Data collection

Study selection followed a systematic two-stage screening process. Initially, two independent reviewers (J.B. and Sh.K.) screened the titles and abstracts according to the predefined eligibility criteria. Any disagreements were resolved through discussion or consultation with a third reviewer (M.Gh.). Subsequently, full-text articles that passed initial screening were thoroughly evaluated for eligibility, with any discrepancies resolved through discussion among reviewers.

Data extraction was performed using a standardized form including first author’s name, publication year, country of origin, study design, sample size, procedure type, intervention details, control group specifications, follow-up duration, number of patients with and without CSF leakage in the intervention and control groups, and adverse events. All extracted data underwent cross-verification to ensure accuracy, with any inconsistencies resolved through consensus among the authors.

### Data synthesis

Meta-analysis was performed to synthesize the data from included studies. Since our outcome was dichotomous (treatment success), the risk ratio and risk difference with their 95% confidence intervals were calculated based on the number of participants with and without treatment success in the intervention and control groups. Prediction intervals were estimated to determine the range in which the effect size of a potential future study is likely to fall, assuming it is randomly selected from the same population as the studies included in our meta-analysis.

Given the heterogeneity among included studies, a random-effects model was applied using the restricted maximum likelihood ratio (REML) method. Heterogeneity was assessed using Cochrane’s Q test I^2^ statistics, with the following categories based on I^2^ values: 0-25% (low), 25-50% (moderate), 50-75% (substantial), and 75-100% (high)^14^. Doi plots, LFK indexes, and trim-and-fill methods were used to assess the small study effect (publication bias). We used meta-regression to assess the role of moderator variables on the overall effect size and to explore the potential source of heterogeneity. Subgroup analyses were not performed since we did not have more than 10 studies in our meta-analysis. For sensitivity analysis, a leave-one-out method was used to evaluate the impact of each study on the combined effect.

All statistical analyses were performed using STATA software, version 17. The Cochrane Risk of Bias 2 (RoB-2) traffic light and summary plots were visualized via R-4.4.2 (R Foundation for Statistical Computing, Vienna, Austria, 2021) robvis package. A two-sided P value of <0.05 was considered statistically significant, except for small study effect tests, where a two-sided P value of <0.1 was considered significant.

### Quality assessment

The quality of studies and potential risk of bias of included studies were independently assessed by two authors (J.B. and Sh.K.), with disagreements resolved through consensus. Multiple validated assessment tools were employed based on study design: the Cochrane Risk of Bias 2 (RoB-2) for randomized trials^15^, the Risk of Bias in Nonrandomized Studies of Interventions (ROBINS-I) for non-randomized studies^15,16^, and the Joanna Briggs Institute (JBI) critical appraisal checklist for observational studies^17,18^. Additionally, the overall certainty of evidence was evaluated using the GRADE (Grading of Recommendations, Assessment, Development, and Evaluation) framework^19^. GRADE assesses the quality of evidence across several domains: risk of bias, inconsistency, indirectness, imprecision, and publication bias. Based on these, evidence quality will be classified as high, moderate, low, or very low.

## Results

### Study selection

The initial database search yielded 1,210 results. Following duplicate removal, 584 unique articles were left for screening. Based on the title and abstract review, 551 articles were excluded, leaving 33 articles for full-text assessment. After detailed evaluation, 11 studies were included, of which five met the requirements for quantitative synthesis. The study selection process is illustrated in the PRISMA flow diagram (Figure 1).

**Figure 1.**
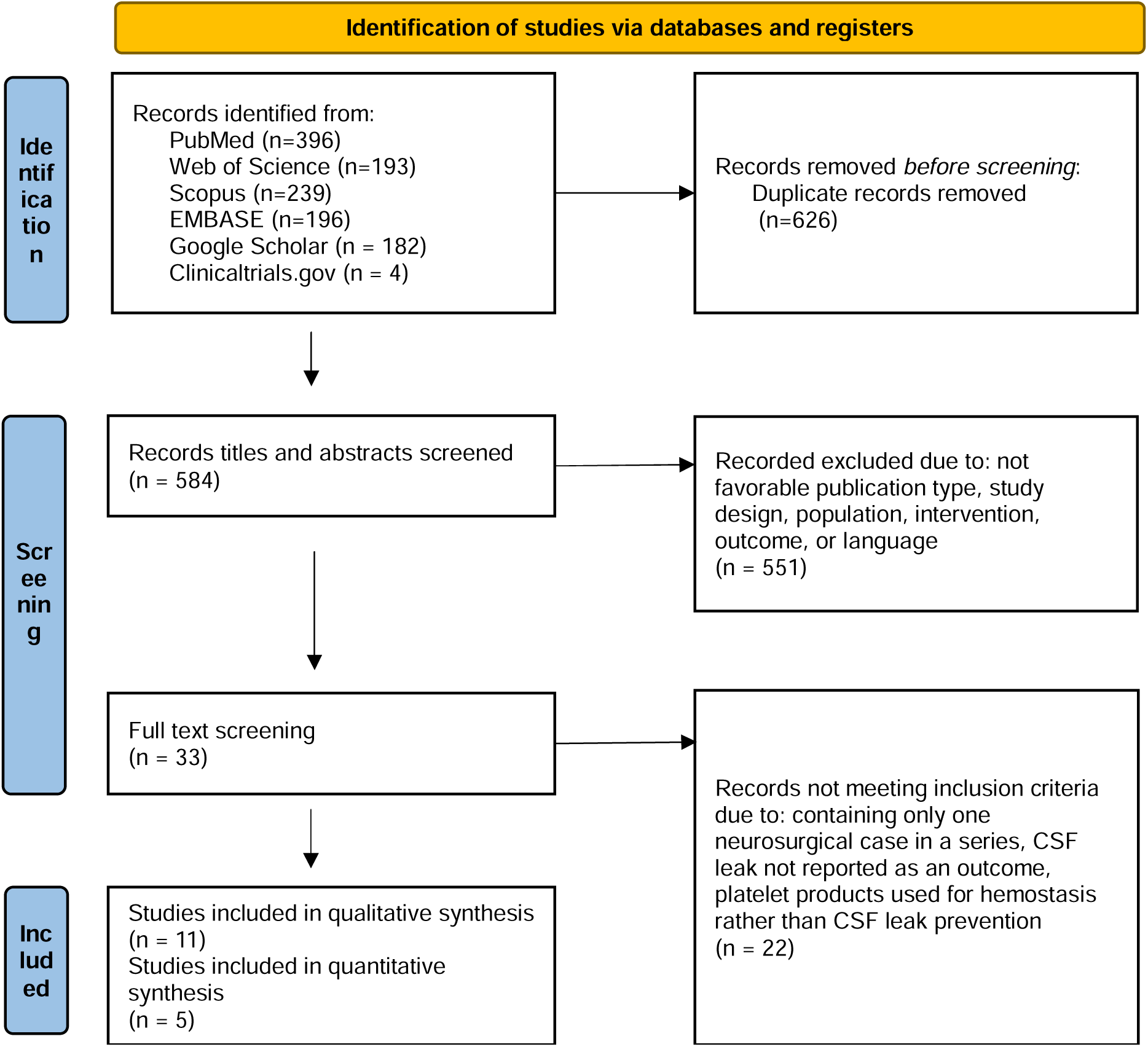
PRISMA flow diagram

### Study characteristics and demographics

This systematic review included 11 studies published between 2017 and 2024, with 5 studies eligible for meta-analysis^8,20–23^. The studies originated from diverse geographical locations: three from Egypt, two from Belgium, and one each from the USA, Argentina, Chile, Turkey, India, and Germany (Table 1). Study designs varied, including four randomized controlled trials, two retrospective studies, three prospective studies, one retrospective cohort study, and one case series. Among the 11 studies, only 6 included control groups^8,20–24^. The total study population encompassed 896 patients, with 543 receiving platelet-rich products. The studies showed a predominance of female patients. The participants in all studies were adults, except for Arabaci et al. which focused on meningocele repair in infants^8^.

**Table 1.** Characteristics of the included studies.

| Author, year | Country | Study design | No. of patients (M/F) | Type of surgery | Intervention/Control | Follow-up duration | Adverse effects | Outcome |
| --- | --- | --- | --- | --- | --- | --- | --- | --- |
| Soldatova (2017)<sup>31</sup> | USA | Retrospective pilot study | 47 (22/25) | Endoscopic endonasal approach for sellar, parasellar, and suprasellar lesions resection | Multilayer reconstruction with L-PRF membranes | 21 days | NR | Postoperative CSF leak occurred in 8.5% (4/47) patients requiring surgical repair<br>1 patient had recurrent CSF leak requiring 3 repairs<br>Improvement in crusting score seen in 42% (17/41) patients |
| Khafagy (2018)<sup>21</sup> | Egypt | RCT | 40 (5/35) | Endoscopic endonasal repair of spontaneous CSF leaks | Intervention: L-PRF as an adjunct layer to standard repair<br>Control: standard multilayer repair with soft tissue graft | 6.8 ± 10.3 months | Persistent headache (50%)<br>Temporary hyposmia (40%)<br>Persistent anosmia (10%)<br>Septal perforation (5%)<br>Posterior meningitis (2.5%) | Failure of repair occurred in 5% intervention group and 15% control group<br>Increased ICP correlated with increased CSF leak (p<0.05)<br>Overall success rate was 95% in intervention group and 85% in control group<br>Revision surgery was successful in all cases of recurrence<br>Difference in recurrence between the 2 groups was found to be statistically insignificant (P value = 0.28). |
| Rasmussen (2018)<sup>32</sup> | Argentina | Prospective pilot study | 12 (7/5) | Endoscopic endonasal transsphenoidal approach to sellar region | L-PRF membrane | 30 days | None | No CSF leak reported in any patient<br>90% mucosal defect restitution |
| Theys (2018)<sup>39</sup> | Belgium | Retrospective feasibility study | 44 (19/25) | Endoscopic transsphenoidal surgery<br>Craniotomy<br>Spine surgery | L-PRF plugs or membranes | 3 months (3-43 weeks) | None | Postoperative CSF leak occurred in only 7% (3/44 patients) of cases where L-PRF was used (2 endoscopic transsphenoidal and 1 spinal CSF leak repair) |
| Ibrahim (2022)<sup>20</sup> | Egypt | Prospective controlled study | 42 (6/36) | Endoscopic endonasal repair of spontaneous CSF leaks | Intervention: two layers of L-PRF membrane + fascia lata +mucosal graft<br>Control: fascia lata + mucosal graft | 12 months | L-PRF vs. control group:<br>Meningitis: 0 vs. 1<br>Headache: 9 vs. 8<br>Severe crusting: 5 vs. 12 | CSF leak recurrence in 1 patient from each group |
| Constanzo (2022)<sup>40</sup> | Chile | Retrospective cohort | 242 (114/128) | Endoscopic endonasal approach for resection of pituitary adenomas and other skull base lesions | Intervention: L-PRF as an adjunct to standard closure protocol<br>Control: standard closure protocol | 6 months minimum | Postoperative meningitis in 3 patients in L-PRF group and 1 patient in control group | Overall postoperative CSF leak rate: 7% (Control) vs. 2% (L-PRF group), $p=0.054$<br>Postoperative CSF leak in intraoperative leak cases: 2% (L-PRF group) vs. 10% (Control), $p=0.02$<br>Postoperative CSF leak in high-flow intraoperative leaks: 3% (L-PRF group) vs. 11% (Control), $p=0.04$<br>L-PRF reduced postoperative CSF leak by 76% (RR=0.24, 95% CI 0.06-0.87) in intraoperative leaks and 73% (RR=0.27, 95% CI 0.07-0.99) in high-flow leaks<br>Most significant reduction in CSF leak was in non-adenoma cases ($p=0.04$) |
| Arabaci (2023)<sup>8</sup> | Turkey | RCT | 40 (20/20) | Surgical repair of meningocele | Intervention: PRP applied during and after surgery<br>Control: Same surgery without PRP | 3 months minimum | PRP vs. Control group:<br>Meningitis 0 vs. 7<br>Skin necrosis 3 vs. 13<br>Wound dehiscence 3 vs. 7<br>Hydrocephalus 8 vs. 15 | CSF leak in 1 patient (5%) in PRP group vs. 9 patients (45%) in control group |
| Ibrahim (2023)<sup>20</sup> | Egypt | RCT | 48 (9/39) | Endoscopic transnasal repair of spontaneous CSF leak | Intervention: multilayer repair with fat enhanced L-PRF membrane<br>Control: multilayer repair with fascia lata | 12 months | L-PRF vs. control group:<br>Meningitis: 0 vs. 1<br>Headache: 7 vs. 9<br>Tight hematoma: 0 vs. 1 | Early CSF leak occurred in 1 patient in each group |
| Coucke (2024)<sup>23</sup> | Belgium | RCT non-inferiority | 328 (146/182) | Elective cranial surgery | Intervention: L-PRF<br>Control: Commercially available fibrin sealants (mostly TachoSil and Tisseel) | 12 weeks | L-PRF vs. Control group:<br>Infection: 4 vs. 6<br>Hemorrhage: 4 vs. 3<br>Meningitis: 2 vs. 6 | CSF leak occurred in 1 patient in L-PRF group, and 5 patients in control group<br>1 patient in control group needed revision surgery for CSF leak<br>Overall relative risk for CSF leak was 0.20 (95% CI $-\infty$ to 1.02, $p = 0.11$) confirming non-inferiority of L-PRF<br>Cost of L-PRF was significantly lower |
| Yadav (2024)<sup>10</sup> | India | Prospective study | 31 (12/19) | Endoscopic anterior skull base defect repair | L-PRF | 2 years | 1 postoperative meningitis<br>1 required revision surgery due to CSF leak recurrence | 97% success rate in closure of CSF leak (30/31 patients) |
| Shah (2024)<sup>7</sup> | Germ any | Case series | 22 (9/12) | Endoscopic transnasal transsphenoidal surgery for pituitary adenoma resection | three-layer method using solid membranous (s-PRF) and high viscosity injectable (i-PRF) forms of PRF | 12 months | 1 patient (Grade 3 intraoperative leak) developed postoperative leak, successfully treated with a lumbar drain | 95% success rate in preventing postoperative CSF leak (21/22 patients)<br>94% success rate in patients who had an intraoperative CSF leak (16/17 patients) |

**Abbreviations:** CI: confidence interval, CSF: cerebrospinal fluid, ICP: intracranial pressure, iPRF: injectable platelet-rich fibrin, L-PRF: leukocyte-rich platelet fibrin, M/F: male/female, NR: not reported, PRF: platelet-rich fibrin, PRP: platelet-rich plasma, RCT: randomized controlled trial, RD: risk difference, RR: relative risk, s-PRF: solid membranous platelet-rich fibrin

### Surgical procedures and interventions

The majority of procedures (9 out of 11 studies) involved endoscopic endonasal approaches for various skull base pathologies, and the remaining studies examined elective cranial surgery, spine surgery, and meningomyelocele repair (Table 1).

Leukocyte-rich platelet fibrin (L-PRF) emerged as the most common platelet-rich product, used in 9 of the 11 studies. The L-PRF was primarily applied as membranes or plugs, with some studies using multiple layers. One study employed a unique three-layer method combining solid membranous PRF (s-PRF) with high-viscosity injectable PRF (i-PRF)^7^. Arabaci et al. used platelet-rich plasma in liquid form applied during and after surgery^8^. Follow-up duration ranged from 21 days to 2 years, with most studies maintaining follow-up for at least 6 months.

### Clinical outcomes and complications

The primary outcome focused on CSF leak occurrence, with adverse events as secondary outcomes. These outcomes are summarized in Table 1. Platelet-rich products demonstrated favorable results in preventing CSF leaks. In controlled studies, the intervention groups consistently showed lower CSF leak rates compared to control groups.

Complication profiles favored platelet-rich product use in multiple studies. Meningitis occurred less frequently in intervention groups, with several studies reporting zero cases compared to multiple occurrences in control groups. Wound healing complications, particularly in endoscopic skull base surgeries, showed improvement with reduced crusting and better mucosal healing in the intervention group^20^. While headache rates remained similar between groups^20–22^, other complications such as wound dehiscence and skin necrosis occurred less frequently with platelet-rich products^8^. Notably, no studies reported serious adverse events directly attributed to the use of platelet-rich products.

### Meta-analysis results

Five studies were included in the meta-analysis, with details provided in Table 2. Analysis of these studies revealed trivial to small effect sizes in CSF leak treatment, as illustrated in the forest plots (Figure 2). The relative risk analysis showed a trivial but marginally significant effect (RR: 1.03, 95% CI: 1.00 to 1.06, P = 0.06), with a prediction interval of 0.981 to 1.074. The risk difference analysis demonstrated a small, non-significant effect (RD: 0.07, 95% CI: −0.03 to 0.17, P = 0.18), with a prediction interval of −0.261 to 0.397.

**Figure 2.**
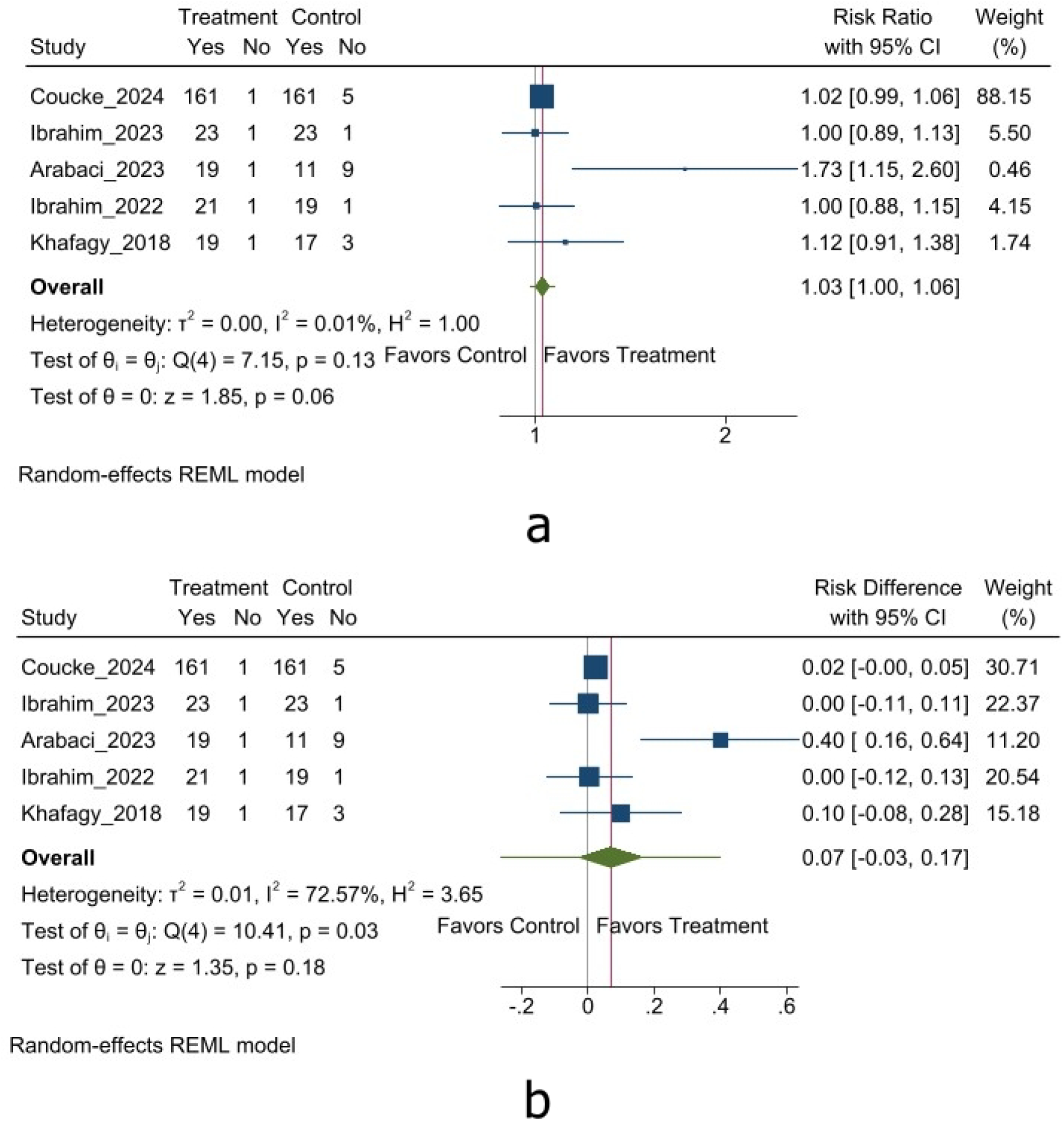
Efficacy of Platelet-Rich Products in Reducing CSF Leaks Following Cranial and Spinal Surgeries: (a)RR, (b)RD

**Table 2.** Characteristics of studies included in the quantitative analysis.

| Author, Year | Number of patients |  | Age (mean ± SD) | Platelet-rich product | CSF leak |  | RR (95% CI) | P-value | RoB |
| --- | --- | --- | --- | --- | --- | --- | --- | --- | --- |
|  | Intervention | Control |  |  | Int | Ctrl |  |  |  |
| Khafagy (2018) | 20 | 20 | 36.9 ± 10.2 | L-PRF | 1 (5%) | 3 (15%) | 1.12 (0.91-1.38) | 0.28 | High |
| Ibrahim (2022) | 22 | 20 | 51 | L-PRF | 1 (4%) | 1 (5%) | 1.00 (0.88-1.15) | 0.47 | High |
| Arabaci (2023) | 20 | 20 | infants | PRP | 1 (5%) | 9 (45%) | 1.73 (1.15-2.6) | 0.003 | High |
| Ibrahim (2023) | 24 | 24 | 41.5 | L-PRF | 1 (4%) | 1 (4%) | 1.00 (0.89-1.13) | 0.49 | High |
| Coucke (2024) | 162 | 166 | 52.2 ± 15.1 | L-PRF | 1 (0%) | 5 (3%) | 1.02 (0.99-1.06) | 0.11 | Low |

**Abbreviations:** CI: confidence interval, L-PRF: leukocyte-platelet-rich fibrin, PRP: platelet-rich plasma, Int: intervention group, Ctrl: control group, RR: risk ratio, RoB: risk of bias

Heterogeneity assessment showed contrasting results between measures. The relative risk calculation exhibited low heterogeneity (I² = 0.01%, τ² = 0.00, P = 0.13), while the risk difference calculation demonstrated substantial heterogeneity (I² = 72.57%, τ² = 0.01, P = 0.03) and should be interpreted cautiously. Subgroup analysis was not methodologically feasible due to the small number of included studies. For evaluating the source of heterogeneity, we used meta-regression, examining the influence of mean age and female participant percentage, which showed no significant impact on the combined effect measure (Figure 3 and Supplementary Table 2).

**Figure 3.**
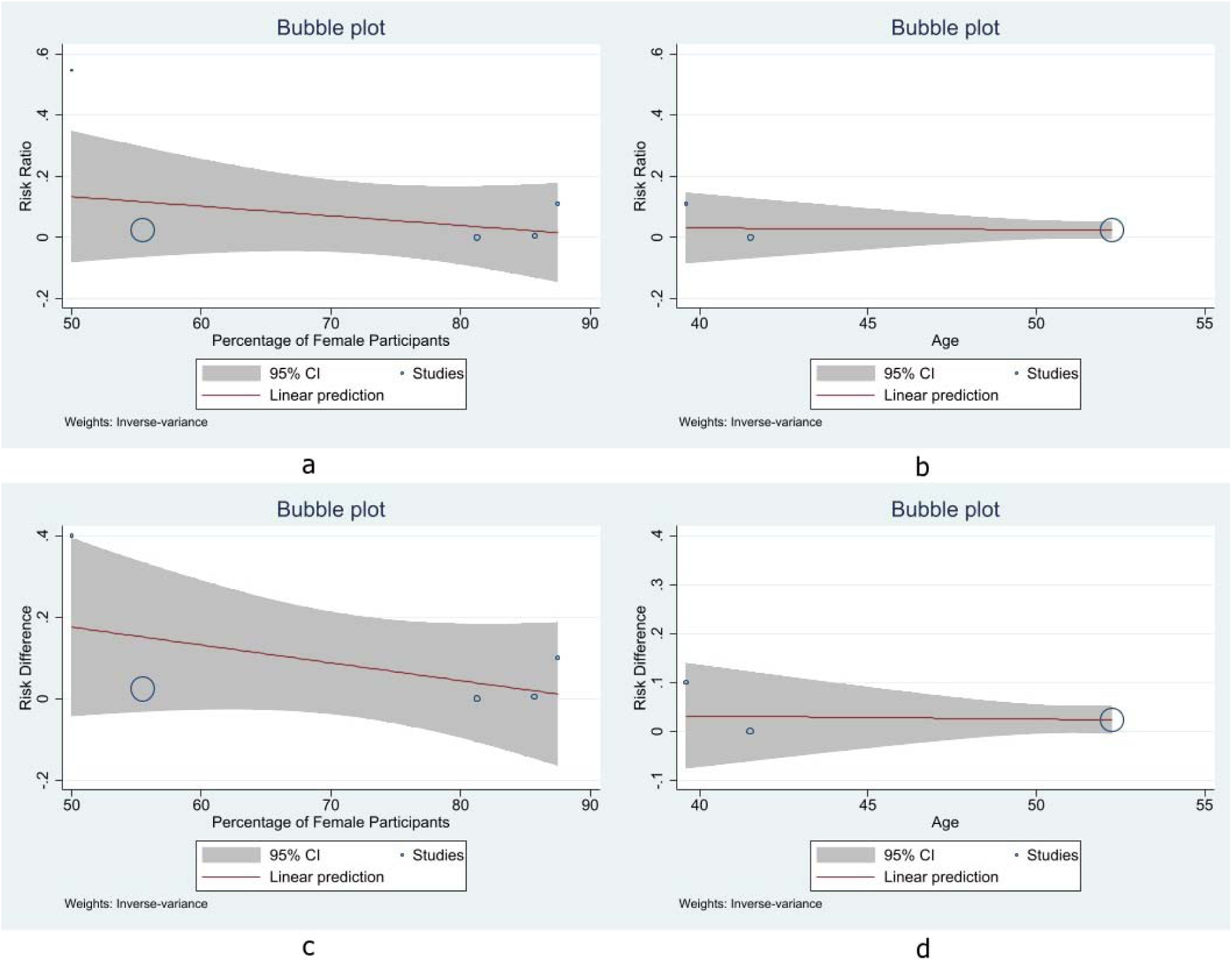
Meta-regression for the Efficacy of Platelet-Rich Products in Reducing CSF Leaks Following Cranial and Spinal Surgeries: (a)effect of percentage of female participants on RR, (b)effect of mean age on RR, (c)effect of percentage of female participants on RD, (d)effect of mean age on RD

### Risk of bias assessment

The risk of bias assessment was conducted using multiple validated tools based on the study design. The Joanna Briggs Institute (JBI) critical appraisal tool was used for observational studies ^17,18^. Although the checklist does not establish a minimum threshold for study inclusion, all studies met acceptable quality standards with no significant risk of bias (Supplementary Tables 3 and 4). The ROBINS-I tool was used in one non-randomized trial, showing moderate overall bias due to potential residual confounding, with low risk in all other domains (Supplementary Table 5). The Cochrane RoB-2 tool was used to assess the risk of bias in randomized trials. Only one study demonstrated a low risk of bias (high quality), while the remaining showed a high risk (low quality). Only Coucke et al. clearly described their randomization process and maintained patient blinding^23^. Documentation of follow-up data proved insufficient across studies, with only one study explicitly reporting loss to follow-up. The absence of registered or shared protocols in most studies prevented the evaluation of protocol deviations and selective reporting of results. Detailed risk of bias assessment is presented in Figure 4.

**Figure 4.**
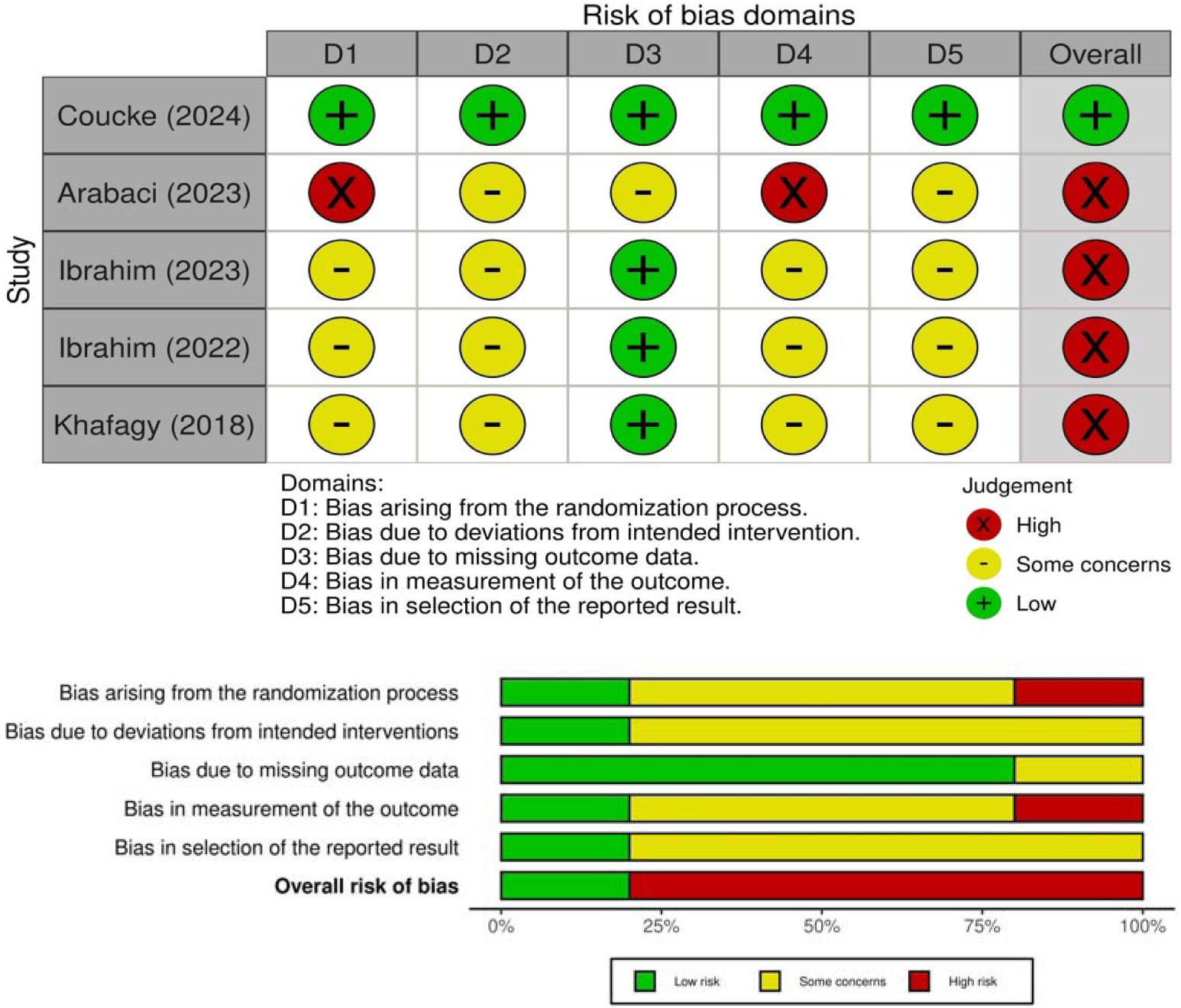
Risk of Bias Evaluation for the Efficacy of Platelet-Rich Products in Reducing CSF Leaks Following Cranial and Spinal Surgeries: (a) Traffic Light Plot, (b)Weighted Bar Plot

### Publication bias and sensitivity analysis

While traditional funnel plot analysis proved unfeasible due to the small number of studies (<10), alternative methods, including Doi plots, LFK indexes, and trim and fill analyses, revealed no considerable small study effect. Results are shown in Figure 5, Supplementary Figure 1, and Table 6.

**Figure 5.**
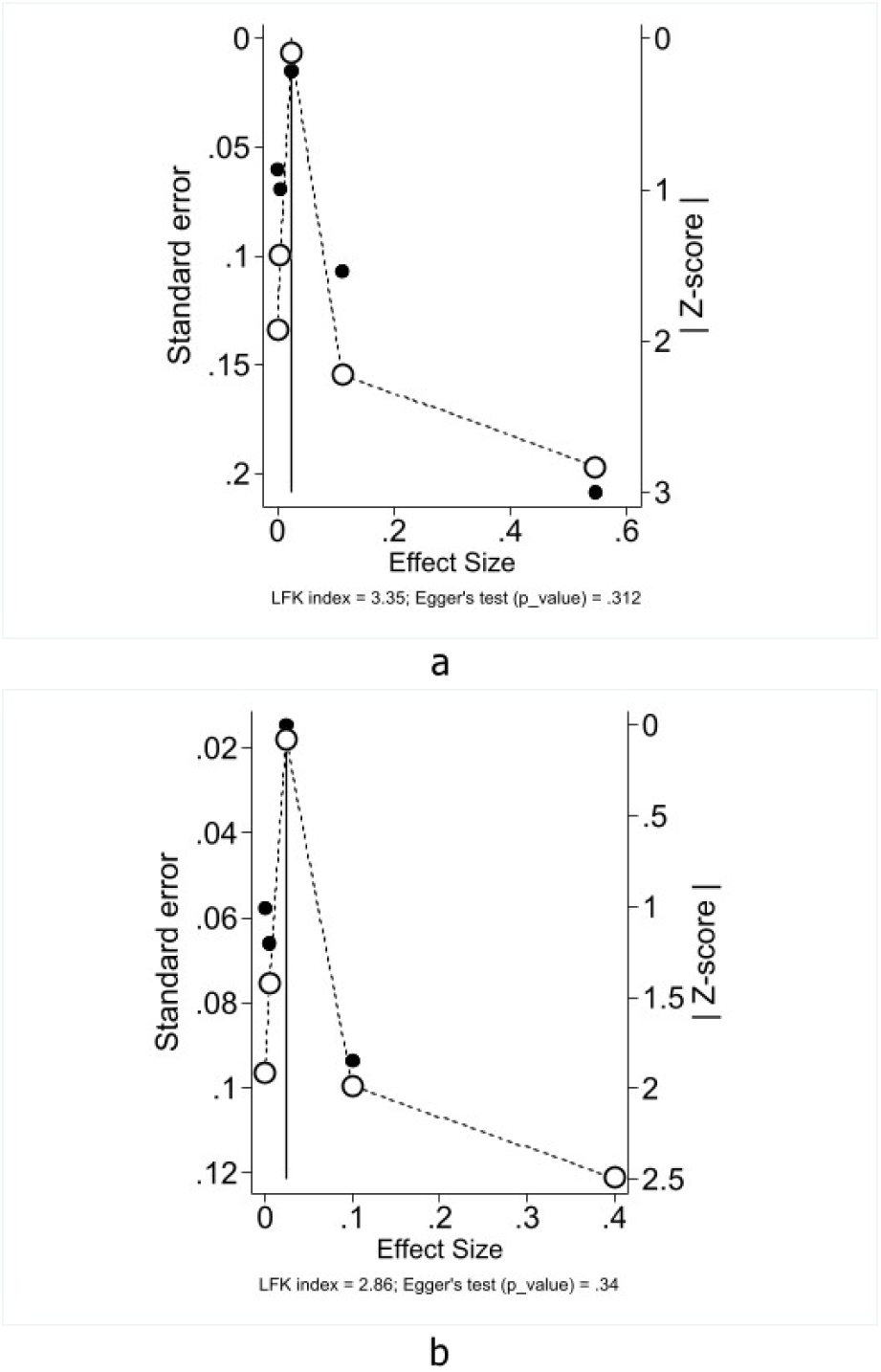
Doi Plots for Small Study Effect (Publication Bias) Assessment for Efficacy of Platelet-Rich Products in Reducing CSF Leaks Following Cranial and Spinal Surgeries: (a) RR, (b) RD

Sensitivity analysis using the leave-one-out method confirmed the robustness of our findings, with no single study substantially impacting the overall results. Results are presented in Figure 6.

**Figure 6.**
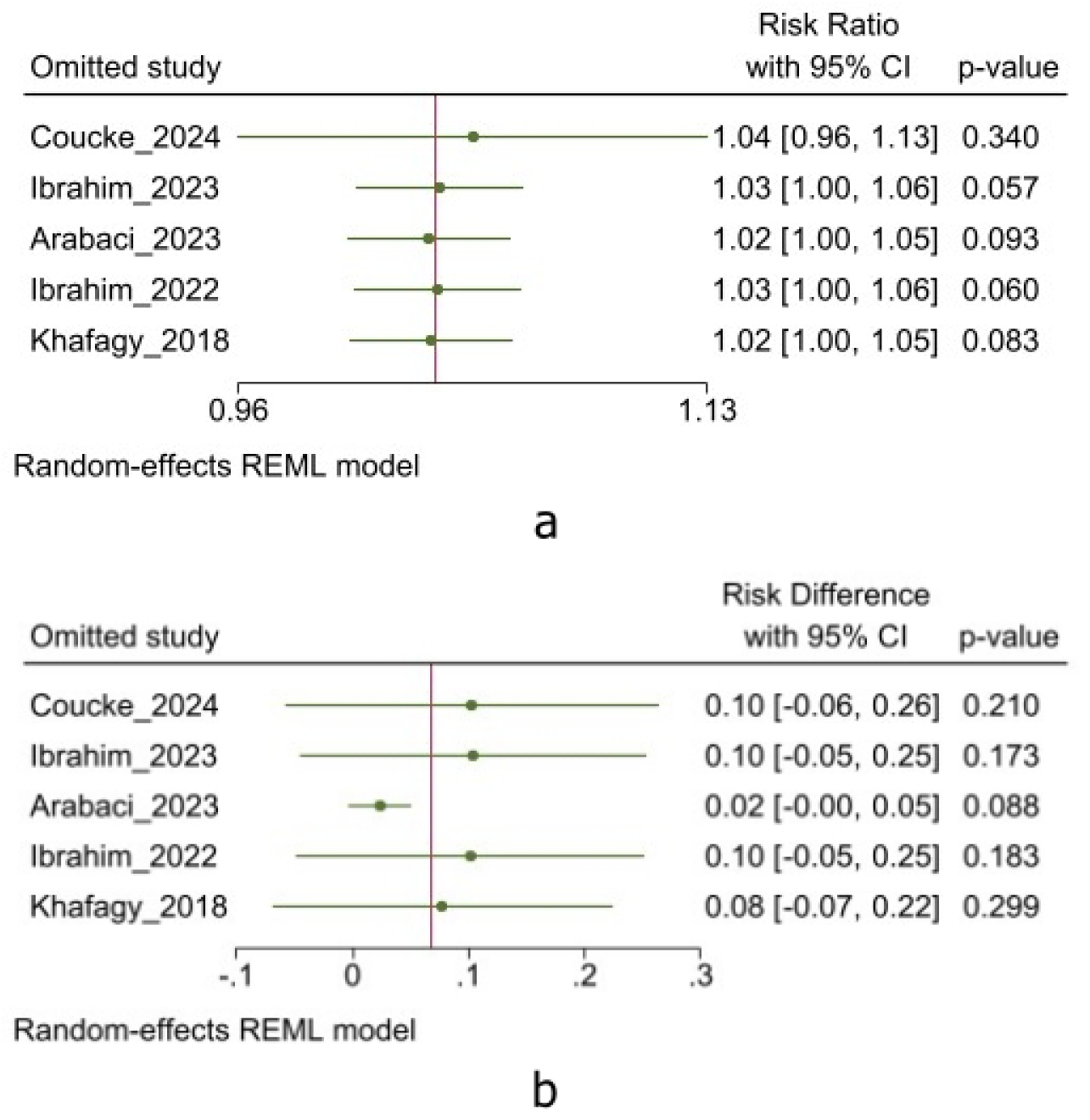
Leave-one-out Sensitivity Analysis Results for Efficacy of Platelet-Rich Products in Reducing CSF Leaks Following Cranial and Spinal Surgeries: (a) Risk Ratio, (b) Risk Difference

### Certainty of evidence

The overall certainty of evidence ranged from low to very low, primarily due to the risk of bias, imprecision, and inconsistency across studies. A detailed GRADE assessment is presented in Supplementary Table 7.

## Discussion

To the best of our knowledge, this systematic review and meta-analysis is the first to evaluate the efficacy of platelet-rich products in reducing cerebrospinal fluid leakage following neurosurgical procedures. We found that platelet-rich products did not significantly reduce CSF leak rates. Our quantitative synthesis of five controlled studies showed a relative risk of 1.03 (95% CI: 1.00 to 1.06, P = 0.06) and a risk difference of 0.07 (95% CI: −0.03 to 0.17, P = 0.18), suggesting that while platelet-rich products may offer some benefit in reducing CSF leak rates, this effect does not reach statistical significance. However, our qualitative analysis showed that studies consistently demonstrated lower rates of CSF leakage in groups treated with platelet-rich products than in controls. This warrants further investigation through well-designed clinical trials with larger populations to better understand the potential role of these products in postoperative CSF leak prevention.

Various neurosurgical interventions necessitate dural opening, exposing patients to the risk of CSF leakage - a major complication in neurosurgery^1^. This may present as rhinorrhea, otorrhea, or leakage through the surgical incision site^1^. CSF leakage is associated with increased infection risk, elevated intracranial pressure, longer hospital stays, and higher mortality rates^25^. Although reported rates vary across the literature, CSF leaks occur more frequently in skull base surgeries and posterior fossa craniotomies compared to spinal and supratentorial craniotomies^2^. Consequently, diverse management strategies have evolved to address this complication, ranging from advanced surgical techniques to innovative biological and synthetic materials. Among these, platelet concentrates have been investigated as promising adjunctive measures for dural repair and CSF leak prevention.

Platelet-rich products have gained widespread attention in different surgical fields such as plastic surgery, otolaryngology, orthopedics, maxillofacial surgery, and neurosurgery^8,26–28^. This is primarily due to their regenerative properties and ease of use^27^. Different platelet-rich products vary in composition because of differences in preparation methods, and this lack of standardization contributes to the inconsistent efficacy seen across clinical studies^29^. Ehrenfest et al. classified these products based on platelet, leukocyte, and fibrin content into pure PRP (P-PRP), leukocyte-rich PRP (L-PRP), pure platelet-rich fibrin (P-PRF), and leukocyte-rich PRF (L-PRF)^30^. Among these, L-PRF, a second-generation platelet concentrate, was the most commonly used product in neurosurgical cases.

One preparation method for L-PRF, as described by Soldatova, involves collecting blood in anticoagulant-free tubes and centrifuging it at 2700 rpm for 12–18 minutes (adjusted based on coagulation status), which will activate the coagulation cascade^31^. During this process, a fibrin clot is formed above the red blood cell layer. The clot is extracted and put in an Xpression™ preparation box and compressed to form a 1-mm thick membrane^31^. Within two hours, the L-PRF membrane is ready for application, maintaining its biological potency for tissue regeneration and surgical repair. Since L-PRF is prepared in parallel with surgery, it does not extend the overall operative time, allowing integration into the surgical workflow^7,32^.

The fibrin scaffold embedded with viable autologous cells and concentrated growth factors - such as transforming growth factor-β (TGF-β), platelet-derived growth factor (PDGF), fibroblast growth factor (FGF), hepatocyte growth factor (HGF), and vascular endothelial growth factor (VEGF)-promotes tissue regeneration through enhanced angiogenesis and vasculogenesis^10^. Also, its platelet and leukocyte components provide bactericidal effects ^33^. The autologous nature of these products eliminates immunological concerns and transmission risks^23^.

Intraoperative measures to prevent CSF leakage following neurosurgical procedures rely on multiple layers of repair, including dural closure, bone and soft tissue reconstruction, as well as CSF diversion when necessary, typically using lumbar drains^34^. Complete dural closure is of paramount importance, but when primary closure is impossible due to dural damage or shrinkage, particularly in prolonged surgeries in some anatomical regions, dural grafts become necessary^35^. Dural grafts are available in two categories: biological and synthetic. Biological options include autologous grafts like fascia lata and pericranium, as well as commercial products such as DuraGen^®^ (bovine) and TissuDura^®^ (equine), which are type I collagen matrices derived from Achilles tendons^6^. Synthetic alternatives include expanded polytetrafluoroethylene (Teflon) and polyglycolic acid ^6,36^. These grafts are often reinforced with dural sealants such as TachoSil^®^, which is FDA-approved for dural repair reinforcement, and Tisseel^®^, a widely used two-component fibrin sealant, used off-label for this purpose^23^. In skull base procedures, particularly in high-flow CSF leaks with large defects, vascularized flaps (e.g., nasoseptal, middle or inferior turbinate flaps) and the gasket seal technique (a method using a fascia lata graft secured by a rigid buttress to create a watertight closure) are also used for soft tissue and bone reconstruction^37,38^.

In the following sections, we explore the role of platelet-rich products in each step of CSF leak prevention described across different neurosurgical contexts.

### Endoscopic skull base surgery

Soldatova et al. first used L-PRF in 2016 to prevent CSF leaks in endoscopic endonasal procedures^31^. Their reconstruction techniques consisted of a collagen matrix or L-PRF membrane for the intradural layer, followed by a mucoperiosteal graft or flap for the extradural layer, and eventually, covering the borders of the reconstruction by L-PRF membranes^31^. Other studies have adopted a similar approach, incorporating L-PRF membranes into their reconstruction techniques^10,12,20,21,32,39^. Multiple studies specifically compared patients who received L-PRF membranes to those who underwent the same reconstruction without L-PRF^20,21,24^. While CSF leak rates were lower in the L-PRF group, the difference did not reach statistical significance. In a subsequent study, Ibrahim et al. introduced an innovative approach combining liposuction-harvested fat with L-PRF and thrombin to create a fat-enhanced L-PRF membrane, which served as both intradural and extradural grafting material^22^. In a recent study, Shah et al. developed a comprehensive multi-layer technique incorporating solid PRF and injectable PRF^7^. They placed an intradural layer of solid PRF at the sella floor, followed by injectable PRF as a bioadhesive, topped with a second solid PRF membrane layer, and secured with a hemostatic gelatin sponge^7^. This approach achieved 95% success rate in preventing CSF leaks^7^.

### Cranial surgery

Coucke et al. conducted a non-inferiority study comparing L-PRF membranes and liquid L-PRF glue against TachoSil^®^ and Tisseel^®^ in elective craniotomies ^23^. They placed L-PRF membrane over sutured dura while using L-PRF glue to reinforce dural closure, and in cases involving bone defects, L-PRF was combined with bone fragments to enhance reconstruction^23^. They found L-PRF to be non-inferior with significant cost advantages (€11.41 versus €208.10 for commercial sealants)^23^. Their study showed lower risk ratios for CSF leakage in both infratentorial and supratentorial approaches with L-PRF, though the differences were not statistically significant^23^. The marked cost advantage of L-PRF over commercial dural products, with their comparable efficacy, supports their adoption in cost-sensitive healthcare environments.

Theys et al. examined the use of L-PRF in cranial procedures in 11 cases including 9 microvascular decompressions, one petrotentorial meningioma removal, and one supratentorial skull base surgery^39^. In these cases L-PRF membranes were used as graft, either sutured or placed extradurally to support dural repair^39^. In MVD cases only, they also used commercial sealants to assist Teflon patch placement and reintegrate bone fragments^39^. No complications, such as CSF leaks or surgical site hemorrhage were reported in their cases^39^.

### Spine surgery

Platelet-rich plasma has been applied in various aspects of spine surgery, such as treating discogenic back pain, lumbar disc herniation, spinal cord injuries, ligament injuries, spinal fusion, intervertebral disc regeneration, and preventing epidural fibrosis^29^. However, similar to craniotomies, their application in spinal surgeries to enhance dural repair has been limited. In one study, Theys et al. investigated L-PRF for dural repair in two patients with chronic CSF leaks after multiple failed spine surgeries^39^. In one case, a 3-cm dural defect was sutured with L-PRF, while in the other, L-PRF was layered as an extradural graft to reinforce closure^39^. They also used TachoSil® in both cases for extra support^39^. While the second patient improved, the first experienced CSF leak recurrence^39^. In a different application, Arabaci et al. explored the use of platelet-rich plasma (PRP) in repairing meningomyelocele in infants, applying it directly to the dura and wound edges while also infusing it under the skin flap and along the wound line post-closure^8^. This led to significantly lower rates of CSF leak, meningitis, and hydrocephalus in the PRP group compared to the control^8^.

### Clinical implication

These products offer surgeons a versatile tool that can be adapted to various surgical scenarios - from complex skull base reconstructions to cranial and spinal dural repairs. The autologous nature of these products not only ensures safety but also allows for immediate availability during surgery. Also, their dual mechanism of action - promoting tissue regeneration while providing antimicrobial protection - makes them particularly valuable in preventing both CSF leaks and post-operative infections^33^. The cost-effectiveness and straightforward preparation protocols make these products a practical addition to the neurosurgical armamentarium.

### Limitations

Several limitations should be considered when interpreting our findings. First, the small number of included studies (11 total, with only five suitable for meta-analysis) limited our ability to perform subgroup analyses. Second, the certainty of our evidence is low to very low, indicating the need for future studies with high methodological quality in order. Most studies lacked proper documentation of follow-up data and registered protocols, making it difficult to evaluate protocol deviations and selective reporting. Therefore, we recommend that future researchers ensure their studies are conducted using pre-registered protocols. The heterogeneity assessment showed contrasting results between measures, with substantial heterogeneity in the risk difference calculation requiring cautious interpretation. Furthermore, the varying surgical approaches (endoscopic skull base, craniotomy, spine, and meningomyelocele repair), preparation methods, populations (adults vs. infants), and application techniques across studies make it challenging to establish standardized protocols. While our sensitivity analysis confirmed the robustness of our findings, the overall limitations in study quality and quantity suggest the need for more rigorous, well-designed trials to strengthen the evidence base for platelet-rich products in neurosurgical applications.

## Conclusion

Our systematic review and meta-analysis demonstrated that while these products show favorable trends in preventing CSF leaks and reducing complications, the statistical analysis revealed no significant difference. The intervention groups consistently showed lower rates of complications, especially meningitis and wound healing issues, with no reported serious adverse events. While these findings are encouraging, the overall low to very low quality of evidence necessitates the need for well-designed randomized controlled trials with larger sample sizes to establish the efficacy of these products in neurosurgical procedures. Future studies should focus on standardizing protocols and comparing different techniques to optimize clinical outcomes.

## Acknowledgment

None

## Funding

There are no funds allocated for our project.

## Conflict of interest

We have no conflicts of interest

## Data availability

The datasets used and/or analyzed during the current study are available from the corresponding author upon reasonable request.

## Notes

### Competing Interest Statement

The authors have declared no competing interest.

### Funding Statement

This study did not receive any funding

