## Supplementary material for "Efficacy of platelet-rich products in preventing cerebrospinal fluid leakage following neurosurgical procedures: A systematic review and meta-analysis"

**Supplementary materials**

**Supplementary Table 1** Database search strategy

| Database  (Date search was performed) | Search string | Number of results |
| --- | --- | --- |
| PubMed  (12/4/2024) | ("platelet-rich plasma"[MeSH Terms] OR "platelet rich fibrin"[MeSH Terms] OR "platelet-rich plasma"[Title/Abstract] OR "platelet rich fibrin"[Title/Abstract] OR "leukocyte* and platelet* rich fibrin"[Title/Abstract] OR "L-PRF"[Title/Abstract] OR "PRP"[Title/Abstract] OR "PRF"[Title/Abstract] OR "platelet leukocyte gel"[Title/Abstract] OR "PLG"[Title/Abstract] OR "preparation rich in growth factors"[Title/Abstract] OR "PRGF"[Title/Abstract] OR "leukocyte* platelet rich fibrin"[Title/Abstract] OR "platelet concentrate"[Title/Abstract] OR "platelet gel"[Title/Abstract] OR "platelet derivatives"[Title/Abstract] OR "autologous platelet"[Title/Abstract]) AND ("Neurosurgery"[MeSH Terms] OR "Neurosurgical Procedures"[MeSH Terms] OR "Craniotomy"[MeSH Terms] OR "Spine"[MeSH Terms] OR "neurosurger*"[Title/Abstract] OR "neurosurgical procedure*"[Title/Abstract] OR "neurologic surgical procedure*"[Title/Abstract] OR "Craniotomy"[Title/Abstract] OR "craniotomies"[Title/Abstract] OR "craniectom*"[Title/Abstract] OR "cranial surger*"[Title/Abstract] OR "spinal surger*"[Title/Abstract] OR "skull base"[Title/Abstract] OR "spine surger*"[Title/Abstract] OR "spinal fusion"[Title/Abstract] OR "transsphenoidal"[Title/Abstract] OR "trans-sphenoidal"[Title/Abstract] OR "TTS"[Title/Abstract] OR "endonasal"[Title/Abstract] OR "endoscopic endonasal"[Title/Abstract]) | 396 |
| Scopus  (12/4/2024) | TITLE-ABS-KEY("platelet rich plasma" OR "platelet rich fibrin" OR "platelet-rich plasma" OR "platelet-rich fibrin" OR "leukocyte* and platelet* rich fibrin" OR "L-PRF" OR "PRP" OR "PRF" OR "platelet leukocyte gel" OR "PLG" OR "preparation rich in growth factors" OR "PRGF" OR "leukocyte* platelet rich fibrin" OR "platelet concentrate" OR "platelet gel" OR "platelet derivatives" OR "autologous platelet")  AND  TITLE-ABS-KEY(neurosurger* OR "neurosurgical procedure*" OR "neurologic surgical procedure*" OR Craniotomy OR craniotomies OR craniectom* OR "cranial surger*" OR "spinal surger*" OR "skull base" OR "spine surger*" OR "spinal fusion" OR transsphenoidal OR "trans-sphenoidal" OR "TTS" OR endonasal OR "endoscopic endonasal") | 239 |
| Embase  (12/4/2024) | #1 'platelet rich plasma'/exp OR 'platelet rich fibrin'/exp OR #1 ('platelet rich plasma':ti,ab OR 'platelet rich fibrin':ti,ab OR 'platelet-rich plasma':ti,ab OR 'platelet-rich fibrin':ti,ab OR 'leukocyte* and platelet* rich fibrin':ti,ab OR 'L-PRF':ti,ab OR 'PRP':ti,ab OR 'PRF':ti,ab OR 'platelet leukocyte gel':ti,ab OR 'PLG':ti,ab OR 'preparation rich in growth factors':ti,ab OR 'PRGF':ti,ab OR 'leukocyte* platelet rich fibrin':ti,ab OR 'platelet concentrate':ti,ab OR 'platelet gel':ti,ab OR 'platelet derivatives':ti,ab OR 'autologous platelet':ti,ab)  AND  #2 (neurosurger*:ti,ab OR 'neurosurgical procedure*':ti,ab OR 'neurologic surgical procedure*':ti,ab OR Craniotomy:ti,ab OR craniotomies:ti,ab OR craniectom*:ti,ab OR 'cranial surger*':ti,ab OR 'spinal surger*':ti,ab OR 'skull base':ti,ab OR 'spine surger*':ti,ab OR 'spinal fusion':ti,ab OR transsphenoidal:ti,ab OR 'trans-sphenoidal':ti,ab OR 'TTS':ti,ab OR endonasal:ti,ab OR 'endoscopic endonasal':ti,ab) | 196 |
| Web of Science  (12/4/2024) | TS=("platelet rich plasma" OR "platelet rich fibrin" OR "platelet-rich plasma" OR "platelet-rich fibrin" OR "leukocyte* and platelet* rich fibrin" OR "L-PRF" OR "PRP" OR "PRF" OR "platelet leukocyte gel" OR "PLG" OR "preparation rich in growth factors" OR "PRGF" OR "leukocyte* platelet rich fibrin" OR "platelet concentrate" OR "platelet gel" OR "platelet derivatives" OR "autologous platelet")  AND  TS=(neurosurger* OR "neurosurgical procedure*" OR "neurologic surgical procedure*" OR Craniotomy OR craniotomies OR craniectom* OR "cranial surger*" OR "spinal surger*" OR "skull base" OR "spine surger*" OR "spinal fusion" OR transsphenoidal OR "trans-sphenoidal" OR "TTS" OR endonasal OR "endoscopic endonasal") | 193 |
| Google Scholar  (12/4/2024) | ("platelet-rich fibrin" OR "platelet-rich plasma" OR "platelet concentrate" OR "leukocyte-platelet-rich fibrin") AND "cerebrospinal fluid leak" | 182 |
| Clinicaltrials.gov  (12/4/2024) | ("platelet-rich fibrin" OR "platelet-rich plasma" OR "platelet concentrate" OR "leukocyte-platelet-rich fibrin") AND "cerebrospinal fluid leak" | 4 |

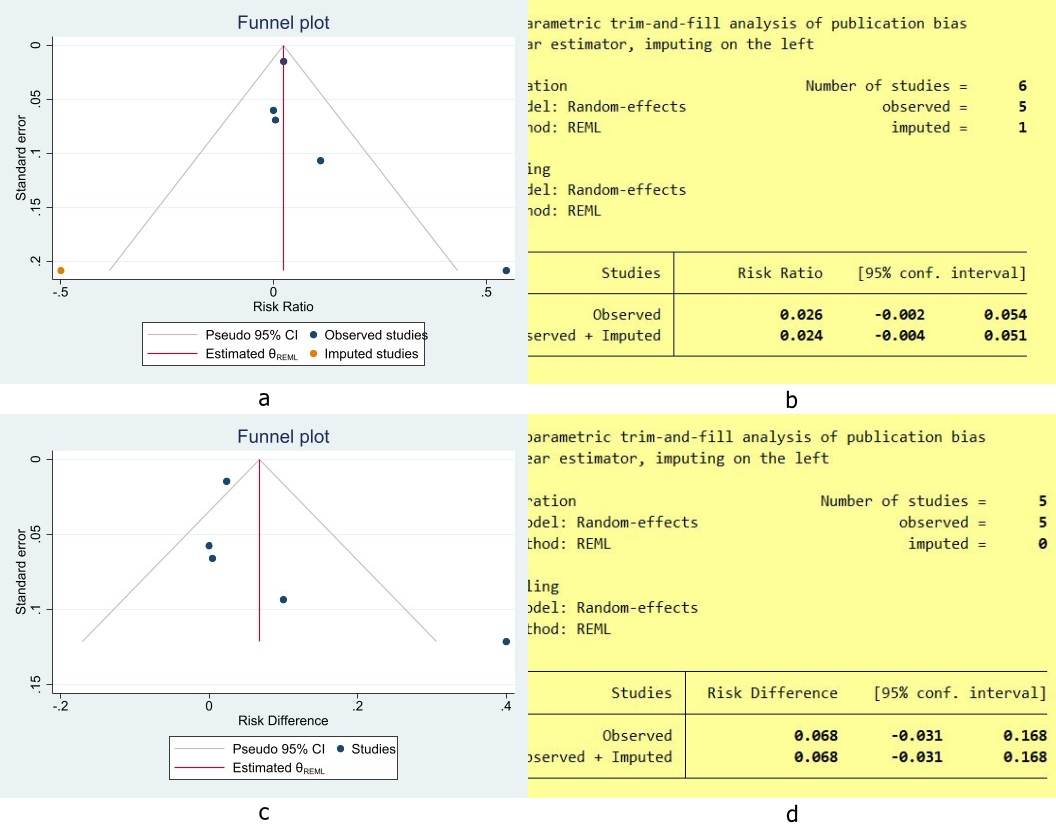

Supplementary Figure 1 Trim-and-Fill Method Results for Efficacy of Platelet-Rich Products) in Reducing CSF Leaks Following Cranial and Spinal Surgeries: (a) trim funnel plots on RR, (b) trim results on RR, (c) trim funnel plots on RD, (d) trim results on RD

**Supplementary Table 2**: Meta-regression Results for Efficacy of Platelet-Rich Products in Reducing CSF Leaks Following Cranial and Spinal Surgeries

| **Variable** | **Coefficient (95% CI)** | **P-Value** | **Residual I2** | **R2** |
| --- | --- | --- | --- | --- |
| **RR** | | | | |
| Female percent | -0.003(-0.011 to 0.005) | 0.433 | 60.09 | 0.00 |
| Mean Age | 0.0005(-0.010 to 0.0089) | 0.912 | 0.00 | 0.00 |
| **RD** | | | | |
| Female percent | -0.004(-0.012 to 0.0037) | 0.289 | 74.35 | 0.00 |
| Mean Age | 0.0006(-0.009 to 0.008) | 0.888 | 0.00 | 74.04 |

Abbreviations: **CI**: confidence interval, **RR**: relative risk, **RD**: risk difference

**Supplementary Table 3** Joanna Briggs Institute critical appraisal checklist for observational studies.

| **Questions** | **Soldatova (2017)** | **Theys (2018)** | **Rasmussen (2018)** | **Yadav (2024)** |
| --- | --- | --- | --- | --- |
| 1. Were the two groups similar and recruited from the same population? | NA | N | NA | N |
| 2. Were the exposures measured similarly to assign people to both exposed and unexposed groups? | NA | Y | NA | NA |
| 3. Was the exposure measured in a valid and reliable way? | Y | Y | Y | Y |
| 4. Were confounding factors identified? | NC | N | Y | N |
| 5. Were strategies to deal with confounding factors stated? | NC | N | N | N |
| 6. Were the groups/participants free of the outcome at the start of the study (or at the moment of exposure)? | Y | Y | Y | Y |
| 7. Were the outcomes measured in a valid and reliable way? | Y | Y | Y | Y |
| 8. Was the follow up time reported and sufficient to be long enough for outcomes to occur? | Y | Y | Y | N |
| 9. Was follow up complete, and if not, were the reasons to loss to follow up described and explored? | Y | Y | Y | NC |
| 10.Were strategies to address incomplete follow up utilized? | NA | NA | N | NC |
| 11. Was appropriate statistical analysis used? | NC | NC | N | NC |
| Overall | Include | Include | Include | Include |

Abbreviations: **Y**: yes, **N**: no, **NA**: not applicable, **NC**: not clear.

**Supplementary Table 4** Joanna Briggs Institute critical appraisal checklist for case series.

| **Questions** | **Shah (2024)** |
| --- | --- |
| 1. Were there clear criteria for inclusion in the case series? | Y |
| 2. Was the condition measured in a standard, reliable way for all participants included in the case series? | Y |
| 3. Were valid methods used for identification of the condition for all participants included in the case series? | Y |
| 4. Did the case series have consecutive inclusion of participants? | Y |
| 5. Did the case series have complete inclusion of participants? | NC |
| 6. Was there clear reporting of the demographics of the participants in the study? | Y |
| 7. Was there clear reporting of clinical information of the participants? | Y |
| 8. Were the outcomes or follow up results of cases clearly reported? | Y |
| 9. Was there clear reporting of the presenting site(s)/clinic(s) demographic information? | Y |
| 10. Was statistical analysis appropriate? | Y |
| Overall appraisal | Include |

Abbreviations: **Y**: yes, **N**: no, **NA**: not applicable, **NC**: not clear.

**Supplementary Table 5** The Risk Of Bias In Non-randomized Studies – of Interventions (ROBINS-I) assessment tool for Costanzo et al. 2022

| **Bias Domain** | **Judgement** | **Support for Judgement** |
| --- | --- | --- |
| 1. Confounding | MODERATE RISK | Controlled for major confounders (age, sex, tumor type, CSF leak grade) but some residual confounding possible due to observational nature |
| 2. Selection of participants | LOW RISK | Clear intervention definition, standardized protocol, data collected at time of surgery |
| 3. Classification of interventions | LOW RISK | Clear eligibility criteria, used prospective database, appropriate exclusions |
| 4. Deviations from intended interventions | LOW RISK | Standardized surgical protocol followed, no evidence of systematic deviations |
| 5. Missing data | LOW RISK | Minimal loss to follow-up (3 patients in Group A), outcomes available for nearly all patients |
| 6. Measurement of outcomes | LOW RISK | Standardized outcome assessment, objective measures used (CSF leak, meningitis) |
| 7. Selection of reported results | LOW RISK | Pre-specified outcomes, appropriate statistical analysis, complete reporting |
| Overall Bias | MODERATE | Main concern is potential residual confounding inherent in observational design |

**Supplementary Table 6** Small study effect (publication bias) results for Efficacy of Platelet-Rich Products in Reducing CSF leaks following Cranial and Spinal Surgeries

| **Effect size** | **LFK index P-value** | **Trim and Fill method results** | |
| --- | --- | --- | --- |
| RR | 0.312 | | Not considerable |
| RD | 0.34 | | Not considerable |

Abbreviations: **RR:** relative risk**, RD**: risk difference

| **Supplementary Table 7** Certainty of evidence according to GRADE approach of Efficacy of Platelet-Rich Products in Reducing CSF leaks following Cranial and Spinal Surgeries | | | | | | | | | | | | |
| --- | --- | --- | --- | --- | --- | --- | --- | --- | --- | --- | --- | --- |
| **Certainty assessment** | | | | | | | **No of patients** | | | | **Effect** | **Certainty** |
| No of studies | Study design | Risk of bias | Inconsistency | Indirectness | Imprecision | Publication bias | | [Participants] | | [Cases] | Relative (95% CI) |  |
| **Risk of CSF leaks** | | | | | | | | | | | | |
| 5 | Trial | Very Serious | None | None | None | None | 498 | | 474 | | RR: 1.03 (1.00 to 1.06) | ⨁⨁◯◯ low |
| **Risk of CSF leaks** | | | | | | | | | | | | |
| 5 | Trial | Very Serious | Very Serious | None | Serious | None | 498 | | 474 | | RD: 0.07 (-0.03 to 0.17) | ⨁◯◯◯ Very low |
| **GRADE:** Grading of Recommendations Assessment, Development, and Evaluation; **CI:** confidence interval; **RR:** risk ratio, **RD:** risk difference | | | | | | | | | | | | |
